# Identifying occupational health inequities in the absence of suitable data: are there inequities in access to adequate bathrooms in U.S. workplaces?

**DOI:** 10.1101/2023.05.11.23289863

**Authors:** Candice Y. Johnson, Kaori Fujishiro

**Author notes:** **Correspondence to:** Candice Johnson, PhD, Division of Occupational and Environmental Medicine, Department of Family Medicine and Community Health, Duke University, 2200 W. Main St, Suite 600, Durham, NC 27705. The findings and conclusions in this report are those of the authors and do not necessarily represent the official position of the National Institute for Occupational Safety and Health, Centers for Disease Control and Prevention.

## Abstract

**Objectives:** Our research questions are often chosen based on the existence of suitable data for analysis or prior research in the area. For new interdisciplinary research areas, such as occupational health equity, suitable data might not yet exist. In this manuscript, we describe how we approached a research project in the absence of suitable data, using the example of identifying inequities in adequate bathrooms in U.S. workplaces.

**Methods:** We created a conceptual model that explained the causation of occupational health inequities, and from this model identified a series of questions that could be answered using separate datasets. Breaking up the analysis into multiple steps allowed us to use multiple data sources and analysis methods, which helped compensate for limitations in each dataset.

**Results:** Using the conceptual model as a guide, we were able to identify jobs that likely have inadequate bathrooms as well as subpopulations potentially at higher risk for inadequate bathrooms. We also identified specific data gaps by reflecting on the challenges we faced in our multi-step analysis.

**Conclusions:** We share our conceptual model and our example analysis to motivate epidemiologists to avoid letting availability of data limit the research questions they pursue.

**What is already known on this topic:** Conducting research in interdisciplinary research areas, such as occupational health equity, can be challenging because suitable data often do not exist.

**What this study adds:** We created a conceptual model that explains the causation of occupational health inequities, which helps conduct analyses with less than optimal data.

**How this study might affect research, practice or policy:** Using this approach allows researchers to combine multiple data sources and analysis methods to answer a single research question, expanding the research questions that can be addressed with existing data.

## INTRODUCTION

The research ideas we pursue are often dictated by data availability or by prior literature showing of the research idea. Sometimes, however, we encounter a topic for which the scope of the problem has not yet been described or no suitable data has yet been collected, and therefore little data exist for future study. Although barriers to conducting research in this situation can be substantial, researchers might be able to create new knowledge from existing data to justify further analyses or data collection.

Finding appropriate data is particularly challenging when researchers examine new topics of interdisciplinary nature, such as occupational health inequities (*i*.*e*., avoidable differences in work-related health outcomes closely linked with social, economic, and/or environmental disadvantages).^1^ Because the close link between work and “social, economic, and/or environmental disadvantages” is not well defined, relevant research fields have not been well integrated, and existing health datasets seldom have adequate data for examining the role of work in creating health inequities across population groups.^2^ This lack of data likely contributes to the slow development of occupational health inequities as a field of investigation.

Here, we use inequities in adequate bathrooms in U.S. workplaces as an example of a potentially important topic for which appropriate data are not readily available. Based on previous small-scale studies and reports, we suspected that workplace bathrooms are inadequate for some workers, but we had no suitable datasets available to directly estimate if inadequate bathrooms is patterned by type of work or worker characteristics.^3–5^ Therefore, we developed a conceptual model and used this model to guide analyses that combined multiple data sources to characterize the potential for inequities in adequate workplace bathrooms. Below we outline in detail our strategy, both conceptual and technical; the decisions we made as we identified potential inequities; and specific data gaps for further investigation.

## ACCESS TO ADEQUATE BATHROOMS AS AN OCCUPATIONAL HEALTH PROBLEM

The U.S. Occupational Safety and Health Administration (OSHA) requires that employers provide bathrooms for their employees and do not prevent employees from using bathrooms as needed; however, many workers report using the bathroom at work less frequently than they want to.^4–8^ Reasons include no bathrooms in the work location (*e*.*g*., transit operators), bathroom deficiencies that discourage their use (*e*.*g*., lack of privacy, uncleanliness, too few toilets, long distances to the bathroom), inability to access available bathrooms (*e*.*g*., bathrooms cannot be used outside of break times, workers too busy, supervisors denying requests for bathroom use), and perceiving bathrooms as unsafe.^3–6,9–11^

Workers who use bathrooms less frequently than needed are more likely to report urinary tract infection and urinary incontinence than those who use bathrooms as needed.^5,12^ Workers with these conditions report lower work productivity and have greater likelihood of work disability, with some workers leaving employment because of their symptoms.^13–15^ These studies indicate that access to workplace bathrooms is an important occupational health issue.

Adequacy of workplace bathrooms may not be uniformly distributed in the U.S. workforce, but we have found no study that has investigated this question. As previous research on workplace bathrooms has focused on women, particularly healthcare workers, inadequate bathrooms in other worker groups or workplaces is not well described.^16^ If we could identify inequities in inadequate workplace bathrooms, we could help explain inequities in health outcomes that might be in part caused by inadequate workplace bathrooms.

## CONCEPTUAL MODEL

We developed a conceptual model that articulates our hypotheses about how inequities in inadequate workplace bathrooms might arise across gender, racial/ethnic, and educational attainment groups (Figure 1). We chose these three worker characteristics because they are the strongest axes of occupational segregation in the U.S. workforce.^17–19^ Our conceptual model consists of two mechanisms of inequity: occupational health inequity and occupational segregation. We posit that occupational health inequities arise because some jobs have characteristics that make bathrooms inadequate, such as lack of plumbing. By identifying such jobs, we would know which occupational groups are at higher risk of inadequate bathrooms.

**Figure 1.**
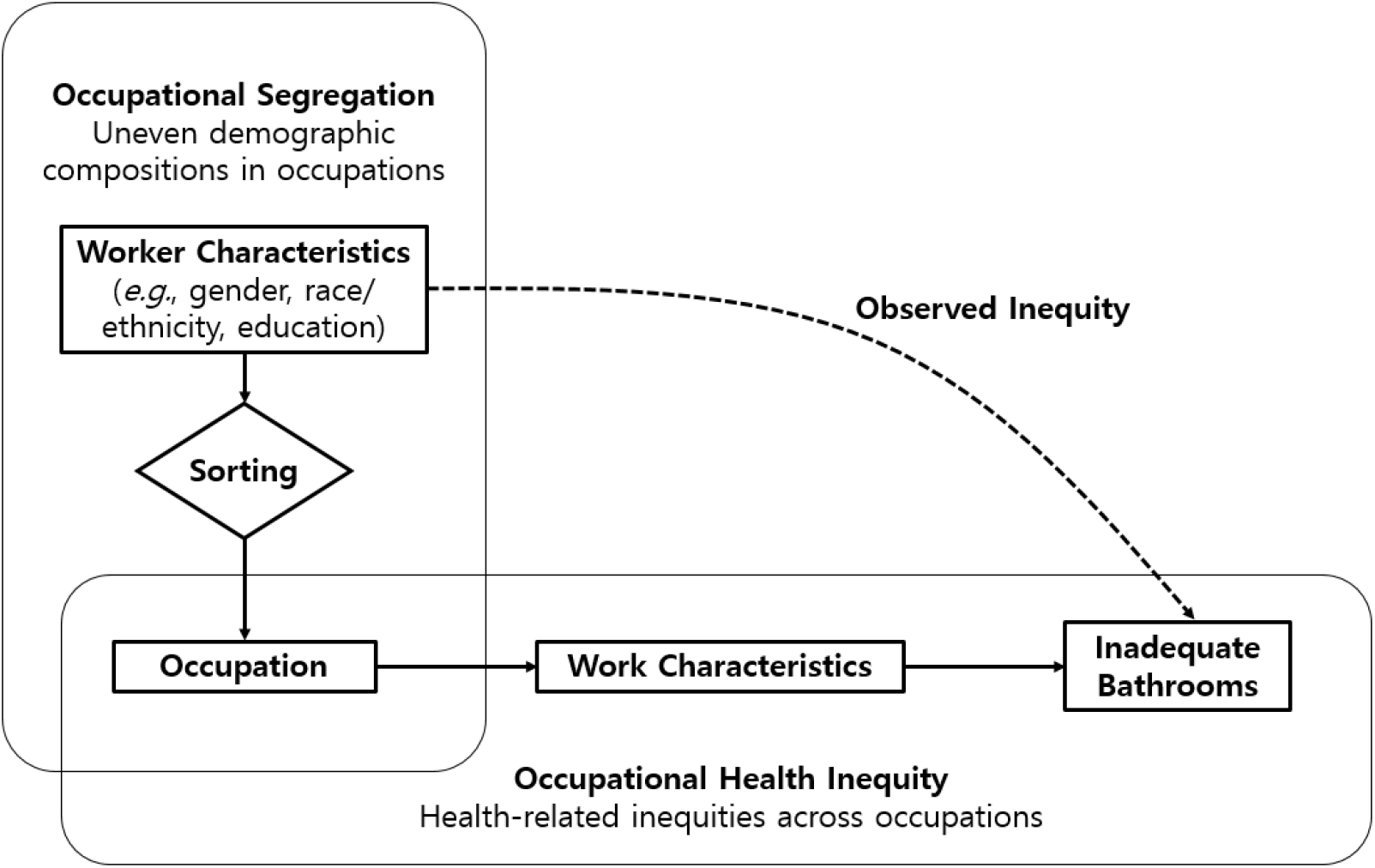
A conceptual model illustrating the mechanisms that cause observed inequities in inadequate workplace bathrooms by worker characteristics: uneven demographic compositions across occupations (*i*.*e*., occupational segregation) and health-related inequities across occupations (*i*.*e*., occupational health inequity).

This difference in risk is an occupational health inequity (*i*.*e*., avoidable differences in work-related health)^1^ because inadequate bathrooms and its related health problems are determined by the person’s job and could be avoided by changing that job’s characteristics. We then posit that various social mechanisms sort people into certain occupations based on demographic characteristics (*i*.*e*., occupational segregation), as observed by uneven compositions in occupation by gender, race/ethnicity, and educational attainment in the U.S. workforce.^19,20^ If certain demographically identified groups are more likely to have occupations with inadequate bathrooms, then occupation explains some of the health inequity related to inadequate bathrooms across demographic characteristics.

## DATA ANALYSIS APPROACH

A conventional approach to identifying inequities might be to directly compare inadequate bathrooms across demographically defined groups (gender, race/ethnicity, educational attainment), as shown in Figure 1 by the direct arrow from worker demographics to inadequate bathrooms. However, we decided against this approach for two reasons. First, our available data did not include a demographically diverse population of workers. Second, and most importantly, that approach fully ignores the cause of inadequate bathrooms. A person’s gender, race/ethnicity, or educational attainment is not the cause of inadequate workplace bathrooms.

Inadequate bathrooms are typically caused by work characteristics like availability of plumbing or work demands. (An important exception is targeted discrimination or harassment. For example, among transgender or non-binary workers, individual characteristics such as gender identity or expression can be a source of discrimination and improper denial of access to workplace bathrooms.^11^) Stated another way, we were not interested in describing how inadequate bathrooms varied by demographic characteristics; rather, we were interested in the potential role of occupation in creating this variation. To investigate the role of occupation in this variation, we needed to identify which jobs offer inadequate bathrooms and who was most likely to hold such jobs.

Our conceptual model guided our analysis. As shown in Figure 2, we conducted a three-step analysis beginning at inadequate bathrooms and working backward through the steps that caused the inequities: work characteristics, occupation, and finally worker characteristics. This approach allowed us to use multiple datasets for three separate analyses instead of finding a single dataset that included all of the information we required.

**Figure 2.**
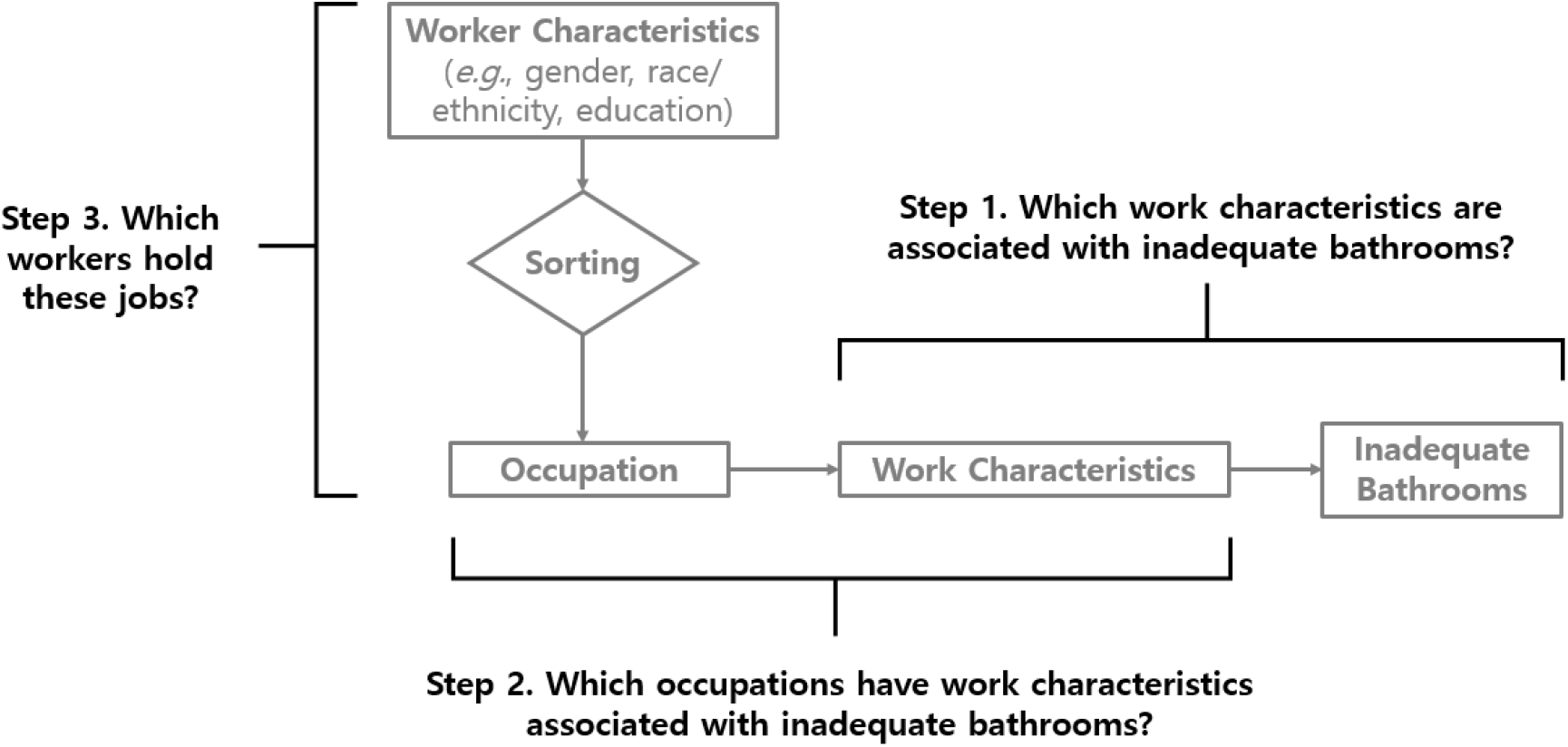
Three steps used to identify potential differences in inadequate bathrooms in the workplace by worker characteristics.

### Step 1: Link work characteristics to inadequate bathrooms

We found one dataset that included both work characteristics and workplace bathrooms: the 2015 American Working Conditions Survey (AWCS), a survey of U.S. workers (see Supplementary Methods for additional details).^21^ Inadequate bathrooms was measured by: “Are you bothered by any of the following in the place you spend most of your working time? … Inadequate toilet facilities.” Respondents could answer yes or no, but no further information was elicited about why bathrooms were inadequate. We then selected questions about work characteristics that might influence adequacy of workplace bathrooms: workplace uncleanliness, working outdoors daily, working in a vehicle daily, work pace set by machine, work pace set by customers, ability to take breaks when needed, and ability to control pace of work. These characteristics or related characteristics have been previously reported in association with inadequate workplace bathrooms.^3,5,6,9,22^

We used Poisson regression with robust standard errors to estimate prevalence ratios (PRs) and 95% confidence intervals (CIs) for associations between work characteristics and inadequate bathrooms, with PR ≥1.5 defined as an association between the characteristic and inadequate workplace bathrooms. This cutpoint was arbitrary, but we had no objective method to determine associations with inadequate bathrooms.

Among the included AWCS respondents (1,019 women and 803 men), 13% of women and 16% of men reported inadequate workplace bathrooms. Of the seven work characteristics, all but ability to control pace of work were associated with inadequate bathrooms (Table 1). The strongest association was with workplace uncleanliness (PR 5.9, 95% CI: 3.8–9.2), followed by working outdoors daily (PR 2.9, 95% CI: 1.8–4.6), working in a vehicle daily (PR 2.2, 95% CI: 1.4–3.7), and work pace set by machines (PR 1.9, 95% CI: 1.2–3.1). Pace set by customers was associated with inadequate bathrooms among men, and ability to take breaks among women.

**Table 1.** Prevalence of inadequate bathrooms by work characteristic and gender—American Working Conditions Survey, 2015.

|  | All Respondents<br>(n = 1,822) |  |  |  | Women<br>(n = 1,019) |  |  |  | Men<br>(n = 803) |  |  |  |
| --- | --- | --- | --- | --- | --- | --- | --- | --- | --- | --- | --- | --- |
|  | N | %<sup>a</sup> | PR | 95% CI | N | %<sup>a</sup> | PR | 95% CI | N | %<sup>a</sup> | PR | 95% CI |
| Workplace cleanliness |  |  |  |  |  |  |  |  |  |  |  |  |
| Not clean | 277 | 48 | 5.9 | 3.8–9.2 | 167 | 44 | 6.6 | 2.9–14.9 | 110 | 51 | 5.5 | 3.3–9.2 |
| Clean | 1,483 | 8 | 1.0 | Reference | 814 | 7 | 1.0 | Reference | 669 | 9 | 1.0 | Reference |
| Missing | 62 |  |  |  | 38 |  |  |  | 24 |  |  |  |
| Works outdoors |  |  |  |  |  |  |  |  |  |  |  |  |
| Daily | 103 | 36 | 2.9 | 1.8–4.6 | 20 | 42 | 3.5 | 1.6–7.7 | 83 | 36 | 2.8 | 1.6–4.8 |
| Less than daily | 1,716 | 12 | 1.0 | Reference | 997 | 12 | 1.0 | Reference | 719 | 13 | 1.0 | Reference |
| Missing | 3 |  |  |  | 2 |  |  |  | 1 |  |  |  |
| Works in a vehicle |  |  |  |  |  |  |  |  |  |  |  |  |
| Daily | 130 | 29 | 2.2 | 1.4–3.7 | 52 | 30 | 2.5 | 1.2–5.3 | 78 | 29 | 2.0 | 1.0–3.9 |
| Less than daily | 1,688 | 13 | 1.0 | Reference | 965 | 12 | 1.0 | Reference | 723 | 14 | 1.0 | Reference |
| Missing | 4 |  |  |  | 2 |  |  |  | 2 |  |  |  |
| Pace set by machine |  |  |  |  |  |  |  |  |  |  |  |  |
| Yes | 334 | 23 | 1.9 | 1.2–3.1 | 187 | 21 | 2.0 | 0.8–4.6 | 147 | 25 | 1.9 | 1.1–3.4 |
| No | 1,488 | 12 | 1.0 | Reference | 832 | 10 | 1.00 | Reference | 656 | 13 | 1.0 | Reference |
| Missing | 0 |  |  |  | 0 |  |  |  | 0 |  |  |  |
| Pace set by customers |  |  |  |  |  |  |  |  |  |  |  |  |
| Yes | 1,395 | 16 | 1.6 | 1.0–2.6 | 783 | 14 | 1.3 | 0.6–2.6 | 612 | 18 | 1.9 | 1.0–3.7 |
| No | 427 | 10 | 1.0 | Reference | 236 | 11 | 1.0 | Reference | 191 | 10 | 1.0 | Reference |
| Missing | 0 |  |  |  | 0 |  |  |  | 0 |  |  |  |
| Can take breaks |  |  |  |  |  |  |  |  |  |  |  |  |
| Not usually | 711 | 18 | 1.5 | 1.0–2.3 | 443 | 17 | 2.1 | 1.1–3.9 | 268 | 18 | 1.2 | 0.7–2.2 |
| Usually | 1,109 | 12 | 1.0 | Reference | 576 | 8 | 1.0 | Reference | 533 | 14 | 1.0 | Reference |
| Missing | 2 |  |  |  | 0 |  |  |  | 2 |  |  |  |
| Can control pace of work |  |  |  |  |  |  |  |  |  |  |  |  |
| No | 370 | 16 | 1.1 | 0.7–1.7 | 224 | 15 | 1.2 | 0.6–2.4 | 146 | 16 | 1.0 | 0.5–2.0 |
| Yes | 1,451 | 15 | 1.0 | Reference | 795 | 12 | 1.0 | Reference | 656 | 16 | 1.0 | Reference |
| Missing | 1 |  |  |  | 0 |  |  |  | 1 |  |  |  |

Abbreviations: CI, confidence interval; PR: prevalence ratio.
<sup>a</sup> Prevalence of inadequate bathrooms by work characteristic (row percent).

### Reflections on Step 1

The objective in Step 1 was to identify work characteristics linked to inadequate bathrooms. AWCS included questions on a variety of work characteristics and inadequate bathrooms. If we did not have this data source, we could have instead relied upon secondary sources (e.g., literature) to identify work characteristics potentially associated with inadequate bathrooms. However, because we had access to these data, we could identify which work characteristics were most strongly associated.

This analysis revealed limitations. First, although we identified seven work characteristics to investigate, other important work characteristics might not have been included in AWCS. Second, AWCS collected no information on why bathrooms were inadequate, information that would allowed us to match specific work characteristics to specific causes of inadequate bathrooms to better inform strategies for improving workplace bathrooms. Some causes of inadequate bathrooms (*e*.*g*., availability and accessibility) are covered by OSHA regulations and can be enforced through inspections and citations, whereas others (*e*.*g*., cleanliness) would likely require other approaches, such as pressure from workers or labor unions.^22^ Third, the AWCS sample size was small relative to the large number of jobs in the U.S. workforce, meaning we could not focus on specific occupational groups or examine differences between them. Supplementary Table 1 shows the jobs in the study population after collapsing them into 23 broad Standard Occupational Classification (SOC) groups; some groups had few or no respondents. Finally, the extent to which the AWCS population is representative of the U.S. workforce is unknown. We therefore used AWCS data as a convenience sample of workers who answered a survey question on bathroom access.

### Step 2: Identify occupations with work characteristics indicating inadequate bathrooms

#### a. O*NET

We used the Occupational Information Network (O*NET, version 24.0) to link occupations with work characteristics.^23^ Of the six work characteristics identified in Step 1, three were included in O*NET: working outdoors, working in a vehicle, and work pace set by machine.

O*NET included information for each job on (i) how often it required workers to be outdoors exposed to weather and outdoors under cover (five categories ranging from never to every day), (ii) how often the job required workers to be in an enclosed vehicle and in an open vehicle (five categories ranging from never to every day), and (iii) the degree to which a work pace determined by speed of equipment was important to each job (five categories ranging from “not important at all” to “extremely important”). O*NET converted the five response categories to a score ranging from 0–100, with higher scores indicating more frequency or importance. For working outdoors and working in a vehicle, we used the highest of the two scores (outdoors exposed to weather/outdoors under cover, enclosed vehicle/open vehicle) as the measure. We sorted jobs by O*NET score and defined jobs with scores ≥75^th^ percentile as having that work characteristic.

Among the 967 jobs with O*NET estimates, 70 (7%) jobs had all three characteristics associated with inadequate bathrooms (working outdoors, working in a vehicle, pace set by machine), 158 (16%) had two, 202 (21%) had one, and 537 (56%) had none. The 10 jobs with the highest scores are shown in Table 2 as examples of the jobs identified.

**Table 2.** Examples of job titles associated with outdoor work, working in a vehicle, and pace set by machine—O*NET version 24.0.

| Outdoor Work |  | Working in a Vehicle |  | Pace Set by Machine |  |
| --- | --- | --- | --- | --- | --- |
| Job Title | O*NET Score | Job Title | O*NET Score | Job Title | O*NET Score |
| Sailors and Marine Oilers | 100 | Police Patrol Officers | 100 | Extruding and Drawing Machine Setters, Operators, and Tenders, Metal and Plastic | 95.2 |
| Railroad Brake, Signal, and Switch Operators | 100 | Refuse and Recyclable Material Collectors | 99.6 | Tire Builders | 92.6 |
| Roustabouts, Oil and Gas | 100 | Fish and Game Wardens | 99.6 | Roof Bolters, Mining | 92.0 |
| Service Unit Operators, Oil, Gas, and Mining | 100 | Wellhead Pumpers | 99.4 | Packaging and Filling Machine Operators and Tenders | 92.0 |
| Pipelayers | 100 | Pest Control Workers | 99.0 | Conveyor Operators and Tenders | 91.6 |
| Meter Readers, Utilities | 100 | Transit and Railroad Police | 98.6 | Computer-Controlled Machine Tool Operators, Metal and Plastic | 89.2 |
| Derrick Operators, Oil and Gas | 99.8 | Fire Inspectors | 97.6 | Logging Equipment Operators | 89.0 |
| Crossing Guards | 99.8 | Logging Equipment Operators | 97.4 | Mine Cutting and Channeling Machine Operators | 88.6 |
| Refuse and Recyclable Material Collectors | 99.6 | Airfield Operations Specialists | 96.8 | Chemical Plant and System Operators | 88.2 |
| Roofers | 99.6 | Telecommunications Line Installers and Repairers | 96.0 | Metal-Refining Furnace Operators and Tenders | 88.2 |

#### b. Revisiting AWCS data

The remaining three work characteristics—uncleanliness, ability to take breaks, and pace set by customers—were unavailable in O*NET, and we found no other dataset linking jobs to these characteristics. We attempted to use AWCS data for this analysis, but there were so few people in each job that we could not identify specific jobs associated with each work characteristic.

##### Reflections on Step 2

We had mixed success. Half of our analysis was a dead-end, and half could proceed to Step 3. Population-based occupational health research can be challenging because of the large sample sizes required to obtain a representative sample of the hundreds of jobs in the U.S. workforce. Datasets like O*NET that include information on nearly all jobs are important supplements to population-based studies. We had concerns about moving on to Step 3 with only half of our work characteristics of interest found in O*NET. Given occupational health’s historical focus on the hazards found in male-dominated manufacturing or manual labor jobs, it would not be surprising if the work characteristics included in O*NET were those predominantly found in those jobs.^24^ Because the prevalence of the six working conditions varies in the AWCS dataset by gender, race/ethnicity, or educational attainment (Supplementary Table 2), we entered Step 3 aware that limiting the next phase of our analysis to these three O*NET work characteristics could drive us to preferentially identify demographic groups concentrated in manual labor jobs, such as male workers of color with less than a college education.

### Step 3. Describe workers in jobs with inadequate bathrooms

We obtained the distribution of gender, race/ethnicity, and education by occupation from the 2015–2019 Current Population Survey (CPS)^25^ and successfully linked 417 jobs to O*NET (see Supplementary Methods for further details). We then reported the demographic composition of jobs with 0, 1, 2, or 3 characteristics (working outdoors, working in a vehicle, pace set by machine) (Table 3). These results reveal apparent inequities. As the number of characteristics associated with inadequate bathrooms increases, jobs have fewer women and fewer college-educated workers, and the racial/ethnic composition of jobs changes.

**Table 3.** Demographic composition of jobs with 0, 1, 2, or 3 characteristics associated with inadequate bathrooms—Current Population Survey, 2015–2019.

|  |  | Jobs With 0<br>Characteristics<sup>a</sup><br>(n = 265) | Jobs With 1<br>Characteristic<sup>a</sup><br>(n = 86) | Jobs With 2<br>Characteristics<sup>a</sup><br>(n = 47) | Jobs With 3<br>Characteristics<sup>a</sup><br>(n = 15) |
| --- | --- | --- | --- | --- | --- |
| Gender |  |  |  |  |  |
| % Women | Range | 1.6–97.7 | 0.5-81.8 | 0.0–44.8 | 0.0–28.0 |
|  | Median | 53.9 | 20.4 | 5.8 | 3.1 |
| Educational attainment |  |  |  |  |  |
| % College Degree | Range | 2.7–100 | 1.3–87.2 | 0.0–77.6 | 1.5–76.0 |
|  | Median | 45.2 | 8.7 | 14.6 | 7.5 |
| Race/ethnicity |  |  |  |  |  |
| % NH White | Range | 23.8–94.9 | 29.1–86.8 | 22.0–92.0 | 36.1–86.8 |
|  | Median | 68.8 | 60.4 | 70.7 | 70.6 |
| % NH Black | Range | 0.6–35.4 | 0.0–36.5 | 0.0–27.1 | 0.0–21.2 |
|  | Median | 8.9 | 9.3 | 9.2 | 6.4 |
| % Hispanic or Latino | Range | 0.0–65.8 | 6.5–54.5 | 2.1–76.4 | 5.9–59.2 |
|  | Median | 10.7 | 20.0 | 13.3 | 19.2 |
| % NH Asian or Pacific Islander | Range | 0.0–58.4 | 0.0–14.0 | 0.0–14.7 | 0.0–4.9 |
|  | Median | 5.4 | 3.3 | 2.2 | 0.8 |
| % NH American Indian or Alaska Native | Range | 0.0–6.6 | 0.0–3.8 | 0.0–2.9 | 0.1–3.3 |
|  | Median | 0.5 | 0.6 | 0.5 | 1.0 |
| % NH Multiracial | Range | 0.0–5.5 | 0.0–3.8 | 0.0–3.7 | 0.1–2.3 |
|  | Median | 1.4 | 1.2 | 1.1 | 0.7 |

Abbreviations: NH, non-Hispanic.
<sup>a</sup> Jobs with 0, 1, 2, or 3 of the follow characteristics: working outdoors, working in a vehicle, or pace set by machine.

#### Reflections on Step 3

O*NET-CPS data had a large enough sample size to generate the demographic distributions of workers within each job; we were no longer restricted by the small sample size of AWCS data. However, we entered Step 3 suspecting that we would identify jobs with a high proportion of male workers of color with less than college education, which we did. Nevertheless, it makes sense that these jobs have inadequate workplace bathrooms because they often involve workspaces with no indoor plumbing or working in situations where a work task cannot be left at-will to use the bathroom. What remains to be seen is who works in the jobs with the other three work characteristics: uncleanliness, inability to take breaks, and work pace set by customers. We also note a limitation of O*NET: a single score for each job. Previous research demonstrates that men and women who hold the same job title perform different tasks^26^ and that O*NET scores correlate with self-reported work characteristics more strongly for non-Hispanic white workers than for racial/ethnic minority workers,^27^ raising the possibility that the data might not accurately reflect the working conditions of some worker groups at high risk for inadequate bathrooms.

#### What we learned about inequities in inadequate workplace bathrooms

Although our attempt to identify inequities was only semi-successful, we identified men, workers with less than a college education, and workers in racial/ethnic minority groups as more likely to work in jobs with characteristics indicating inadequate bathrooms. Given that much of the literature on workplace bathrooms focuses on women in healthcare, the experiences of men in occupations that do not require a college degree could be an important addition to the literature. Thus, our exploration with multiple data sources identified under-investigated worker populations who may be at risk for inadequate workplace bathrooms. A limitation of our analysis was examining gender, racial/ethnic group, and educational attainment separately. Occupational segregation occurs across these three characteristics simultaneously, and examining the intersection of these characteristics could be informative in future research.

Our analysis also identified jobs at high risk for inadequate bathrooms (Table 2). These job titles offer potential target populations for interventions to ensure adequate bathrooms. Because we were unable to pursue analyses of unclean workplaces, inability to take breaks, and a work pace set by customers, the jobs associated with these characteristics remain unknown.Because these characteristics have been reported in studies of working women,^5,6,9^ we might be missing jobs more commonly held by women.

Our findings suggest at least three future directions. First, jobs with characteristics that might predispose workers to inadequate bathrooms (e.g., Table 2) could be starting points for identifying workers for future research studies. Employers following OSHA regulations might already ensure bathroom access in these jobs (*e*.*g*., portable toilets, relief workers), and needs assessments among workers in these jobs could identify unmet needs or lax compliance.

Second, studies of health conditions in occupations at lower and higher risk for inadequate bathrooms could provide additional evidence of the health risks of inadequate bathrooms, furthering efforts to prevent these conditions and to reduce potential inequities.^16^ Third, data on workplace cleanliness, ability to take breaks, and work pace set by customers could be useful if included in population-based or occupation-based data sources. A new national survey from the Bureau of Labor Statistics, the Occupational Requirements Survey, will include ability to pause work in its data collection.^28^

## DISCUSSION

Our objective was to explore how to address a research question for which appropriate data do not exist. As an example, we described combining multiple sets of imperfect data to generate preliminary information about occupational health inequities. Difficulties at each step prevented us from coming to firm conclusions, but our attempt highlighted ways to leverage existing information to identify opportunities for future research and gaps in available data.

### Perfect data do not exist

By presenting our effort to investigate inequities in adequate workplace bathrooms, we hope to inspire researchers to overcome the common situation of data availability dictating what research questions we ask. Instead, researchers can leverage existing data, albeit imperfectly, to expand the horizon of research topics we pursue, including issues of occupational health equity. Our conceptual model was central to this effort and echoes calls for theory-driven, data-informed research.^29–31^ By explicitly hypothesizing the causal mechanisms for inequities, we identified three separate but crucial questions along the causal pathway, and were able to use different datasets to answer them. Although each step had limitations, they revealed ideas for future data collection. Starting a research study with a conceptual model, rather than generating a model based on available variables in a single dataset, helps us to see not only what we could do with limited data but also what could be useful for the future.

Another important strength of the conceptual model was that it steered us away from comparing differences in inadequate bathrooms by gender, race/ethnicity, and education, a typical approach if a large, diverse, individual-level dataset on bathroom access existed. Such analysis would have estimated the risk of inadequate workplace bathrooms by gender, race/ethnicity, education, and other individual-level variables. This type of analysis describes who is at higher risk, but does not inform how to reduce the risk or reduce inequities, because it ignores structural-level (*i*.*e*., workplace-level) causes of inequities in resources. For work-related inequities, the workplace—not the individual—is the most likely intervention point, whether it be by workplace regulations or other methods. The use of our model guided the analysis to inform workplace-based intervention possibilities.

As we reflected upon each step of the analysis, we identified future directions to improve occupational health inequity research more broadly, beyond our example of workplace bathrooms. First, incorporating information on work into population-based epidemiologic studies or social factors into occupational health studies would facilitate understanding a greater array of health inequities. Although work is both a social and structural determinant of health, it is rarely included in epidemiologic studies outside of occupational epidemiology.^2,32^ Studies that include standardized industry and occupation codes and information on work characteristics would allow researchers to understand structural-level health inequities caused by differences in quality of work (*e*.*g*., wages, access to employer-sponsored health insurance, availability of paid leave and retirement benefits). As illustrated in our conceptual model, these differences by occupation–coupled with occupational segregation–are mechanisms by which health inequities can arise. Thus, information on quality of work is crucial to promote research on work as a structural determinant of health and health equity. Second, it is important to include unconventional exposures in occupational epidemiology studies, such as inadequate bathroom access, uncleanliness, and inability to take breaks. Labor unions or employee groups can be valuable sources of information about workplace conditions of concern. These unconventional exposures could be first explored in small-scale studies, which on their own may not offer generalizable knowledge. However, with the approach we demonstrated—use of the conceptual model, multi-step analyses, and careful examinations of data limitations— occupational epidemiologists could use even limited data to address complex emerging issues.

We encourage researchers to consider which research questions are *not* being asked but are worth asking, and how to pursue research to answer them. Even if suitable data do not exist, epidemiologists can advance knowledge by articulating assumptions, maximizing use of fragmented data, and drawing conclusions with appropriate caution. What data are collected and what are not—what is recognized as important for health and what is not—is decided in specific historical and social contexts, which can be changed. Epidemiologists can advance the field so that new problems are brought to light, specific data paucities are identified, and new solutions for current public health problems are found. Data availability does not have to limit our pursuit of important research questions.

## Data Availability

All data produced are available online at: the American Working Conditions Survey (https://www.rand.org/pubs/tools/TL269.html), the Current Population Survey (https://cps.ipums.org/cps/), and O*NET (https://www.onetcenter.org/).

https://cps.ipums.org/cps/

https://www.onetcenter.org/

