## Supplemental Materials for "Identifying occupational health inequities in the absence of suitable data: are there inequities in access to adequate bathrooms in U.S. workplaces?"

### SUPPLEMENTARY MATERIALS

#### METHODS

The Duke University Health System Institutional Review Board determined that this study was exempt from continuing review.

##### Step 1: Link work characteristics to inadequate bathrooms

AWCS was an internet-based survey fielded between July and October 2015 to members of the American Life Panel, a sample of 5,000 households in the 50 U.S. states, the District of Columbia, and Puerto Rico who are regularly invited and paid to complete online surveys on a variety of topics. Participants without computers or internet access received this equipment to allow their participation. 3,131 members of the American Life Panel participated in AWCS, a 64% participation rate among eligible American Life Panel members.

AWCS included questions on work and working conditions related to the respondent's main paid job. Of the 3,131 AWCS respondents, we excluded 1,019 who were not currently working and 41 who worked less than one hour per week, leaving 2,071 eligible workers. Of those, we excluded 249 (12%) who did not answer the question about inadequate bathrooms.

We controlled for no covariates in our models because the purpose of the model was simply to provide a standardized descriptive measure of association across variables. We stratified our analyses by gender because of strong occupational segregation by gender, gender-related differences in bathroom infrastructure (*e.g.*, urinals), and potential gender-related differences in perceptions of bathroom adequacy (*e.g.*, the conditions and number of stalls).

##### Step 2: Identify occupations with work characteristics indicating inadequate bathrooms

We sorted jobs by each O*NET score and defined jobs with scores in the 75th percentile or above as being associated with that work characteristic. The 75th percentile score was 65.2 for working outdoors, 62.2 for working in a vehicle, and 47.8 for pace set by machine.

**Step 3. Describe workers in jobs with inadequate bathroom access**

We obtained Current Population Survey (CPS) data on worker characteristics through the IPUMS CPS website (https://cps.ipums.org/cps/). We linked CPS worker data to O*NET by job code, crosswalking 2010 Census codes to the 2010 O*NET-SOC codes. Because of differences between Census and SOC codes, heterogeneous job categories that do not receive O*NET scores, and uncommon jobs not represented among CPS respondents, we successfully linked 467 jobs. We then excluded 53 jobs (11%) that had <50 people in the CPS sample to avoid including unstable estimates, leaving 413 jobs in the final analysis. Using the same 75th percentile O*NET scores as in Step 2, there were 15 jobs (4%) with all three of working outdoors, working in a vehicle, and pace set by machine, 47 (11%) with two of these work characteristics, 86 (21%) with one, and 265 (64%) with none of these characteristics.

Supplementary Table 1. Broad occupational groups represented among included respondents, by gender—American Working Conditions Survey, 2015.

| | | Women (n = 1,019) | | Men (n = 803) | |
|---|---|---|---|---|---|
| SOC Code | Occupational Group | N | % | N | % |
| 11 | Management | 129 | 14 | 112 | 15 |
| 13 | Business and Financial Operations | 61 | 7 | 61 | 8 |
| 15 | Computer and Mathematical | 12 | 1 | 45 | 6 |
| 17 | Architecture and Engineering | 6 | 1 | 37 | 5 |
| 19 | Life, Physical, and Social Science | 24 | 3 | 16 | 2 |
| 21 | Community and Social Service | 50 | 5 | 17 | 2 |
| 23 | Legal | 19 | 2 | 19 | 3 |
| 25 | Education, Training, and Library | 118 | 13 | 47 | 6 |
| 27 | Arts, Design, Entertainment, Sports, and Media | 20 | 2 | 31 | 4 |
| 29 | Health Practitioners and Technical | 82 | 9 | 23 | 3 |
| 31 | Healthcare Support | 53 | 6 | 9 | 1 |
| 33 | Protective Service | 11 | 1 | 26 | 4 |
| 35 | Food Preparation and Serving Related | 21 | 3 | 14 | 2 |
| 37 | Building and Grounds Cleaning and Maintenance | 15 | 2 | 16 | 2 |
| 39 | Personal Care and Service | 25 | 3 | 6 | 1 |
| 41 | Sales and Related | 73 | 8 | 66 | 9 |
| 43 | Office and Administrative Support | 176 | 19 | 45 | 8 |

| | | | | | |
|---|---|---|---|---|---|
| 45 | Farming, Fishing, and Forestry | [a] | [a] | [a] | [a] |
| 47 | Construction and Extraction | [a] | [a] | 25 | 3 |
| 49 | Installation, Maintenance, and Repair | [a] | [a] | 31 | 4 |
| 51 | Production | 19 | 2 | 23 | 3 |
| 53 | Transportation and Material Moving | 7 | 1 | 45 | 6 |
| 55 | Military Specific | [a] | [a] | 5 | 1 |
| | Missing | 89 | | 70 | |

Abbreviations: SOC, Standard Occupational Classification (2010).

[a] Result suppressed, <5 respondents.

Supplementary Table 2. Prevalence of work characteristics, by gender, educational attainment, and race/ethnicity—American Working Conditions Survey, 2015.

| | Prevalence (%) | | | | | |
|---|---|---|---|---|---|---|
| | Gender | | Education | | Race/Ethnicity | |
| Working Condition | Women | Men | College | No College | Non-Hispanic White | All Other Groups |
| Working outdoors | 3 | 15 | 4 | 13 | 8 | 12 |
| Working in a vehicle | 7 | 11 | 6 | 11 | 7 | 12 |
| Pace set by machine | 25 | 26 | 12 | 32 | 22 | 32 |
| Unclean workplace | 16 | 17 | 11 | 20 | 16 | 17 |
| Inability to take breaks | 50 | 44 | 31 | 55 | 45 | 50 |
| Pace set by customers | 76 | 78 | 78 | 76 | 82 | 68 |
